# A Review of Interventions for Non-Communicable Diseases in Humanitarian Emergencies in Low-and Middle-Income Countries

**DOI:** 10.1101/2021.12.05.21267308

**Authors:** Rebecca Leff, Anand Selvam, Robyn Bernstein, Lydia Wallace, Alison Hayward, Pooja Agrawal, Denise Hersey, Christine Ngaruiya

## Abstract

**Background:** Low-and middle-income countries (LMICs) not only experience the largest burden of humanitarian emergencies but are also disproportionately affected by non-communicable diseases (NCDs). Interventions addressing NCDs require humanitarian entities to consider complex challenges such as continuity of care, diagnostics, logistics and cost of care for recurrent or expensive treatments, yet primary focus on the topic is lacking. We conducted a systematic review on the effects of humanitarian disasters on NCDs in LMICs with the primary aim of identifying studies on epidemiology, interventions, and treatment. Key interventions were identified and their effects on populations in disaster settings were reviewed.

**Methods:** A systematic search was conducted in MEDLINE, MEDLINE (PubMed, for in-process and non-indexed citations), Social Science Citation Index, and Global Health (EBSCO) for indexed articles published before December 11, 2017. Publications reporting on interventions targeting NCDs during disasters in LMICs were included if they incorporated core intervention components as defined by the United States Department of Health and Human Services. Two separate screeners independently evaluated the titles, abstracts and full text of the eligible articles, with vetting by a third reviewer. Key intervention components including target population, phase of crisis, and measured outcomes among others were extracted into a template and synthesized using a thematic analysis approach. The full systematic review is registered at PROSPERO(CRD42018088769).

**Results:** Of 85 articles eligible for the full systematic review, only seven articles describing interventions met inclusion criteria. Studies focused reporting on the response (n=4) and recovery (n=3) phases of disaster, with no studies reporting on the mitigation or preparedness phases. Successful interventions conducted extensive pre-deployment risk assessments to assess the burden and distinct epidemiology of NCDs amongst affected populations, worked in close cooperation with local health services, assessed individual needs of sub-populations in disaster regions in the response phase, promoted task shifting between humanitarian and development actors, and adopted flexibility in guideline implementation. Training and capacity building of staff were found to be essential elements of successful interventions due to an assessed lack of experience of healthcare workers in disaster settings with NCDs and successfully allowed for incorporation of community health workers.

**Conclusions:** We found only limited interventions designed to address NCDs in humanitarian emergencies, with a particular dearth of studies addressing the mitigation and preparedness phases of humanitarian response. Delivering interventions for NCDs in humanitarian emergencies requires improved collaboration between humanitarian and development actors in addition to improved NCD training and capacity building amongst healthcare workers in disasters settings.

## BACKGROUND

Non-communicable diseases (NCDs) represent the leading cause of mortality worldwide, accounting for 70% of deaths globally [1]. Almost three quarters of NCD-related deaths occur in low and middle-income countries (LMICs), where the steepest increases in morbidity and mortality from NCDs are projected to occur over the coming decades [2, 3]. In Africa alone, deaths due to NCDs are projected to exceed those from communicable, maternal, perinatal, and nutritional diseases combined by 2030 [4].

NCDs often occur in conjunction with humanitarian emergencies in LMICs, generating added challenges for NCD management, as strained healthcare systems are faced with the additional burden of disaster response [5]. Humanitarian emergencies are defined by the World Health Organization (WHO) as “large-scale events that affect populations or societies causing a variety of difficult and distressing consequences that may include massive loss of life, disruption of livelihoods, breakdown of society, forced displacement, and other severe political, economic, social, psychological and spiritual effects [6].” Currently, more people are affected by humanitarian crises —such as armed conflicts, natural disasters, pandemics, and forced displacement — than any period in documented history [7]. The United Nations High Commissioner for Refugees (UNHCR) Global Trends Report estimates that an unprecedented 79.5 million people were displaced from their homes as internally displaced persons (IDPs) or refugees in 2019 - the largest figure ever recorded [8].

Disaster response can be divided into four phases: mitigation, preparedness, response, and recovery [9]. Mitigation describes measures aimed at either preventing or reducing the impact of disasters. Preparedness refers to training and preparation aimed at enhancing the overall capacity and capability of a state or community for events that cannot be prevented. The response phase describes the immediate aftermath of a disaster when disaster response plans are implemented and resources are mobilized. Finally, the recovery phase is comprised of restoration efforts, which occur in parallel with routine operations and activities [9].

Each phase may be prolonged, exemplified by current conflicts in Israel-Palestine, Syria, Yemen, the Democratic Republic of Congo, and countries of the Lake Chad region which are party to protracted conflicts where the response phase has persisted [10]. Protracted crises not only play a role in dictating approaches to NCD management in disasters, but also impact the magnitude of the burden. The increasing prevalence of protracted cries in higher income regions, (e.g. Eastern Mediterranean region (EMRO)) compounded by a drive towards displaced persons residing in urban districts rather than traditional refugee camp sites have increased the likelihood of disaster affected populations having pre-existing NCDs or NCD risk factors [5, 11].

Despite these significant global shifts, preventative interventions, diagnostics, and therapeutic interventions for long-term management and acute complications of NCDs have largely been neglected in humanitarian emergencies [12-14]. It is evident that our approach to health intervention in humanitarian disasters must evolve substantially, yet evidence-based models are lacking [5]. Further research is required to develop cost efficient, effective interventions and models of care to adequately address NCDs in humanitarian emergencies [5].

On this basis, we conducted a systematic review on the effect of humanitarian disasters on NCDs in LMICs assessing epidemiology, interventions, and treatment and found solutions among a limited number of interventions. Key interventions from the review were identified and their effects on populations in various stages of humanitarian crises are described here. Our aim is to guide allocation of resources, future research, and intervention development. The full systematic review is published elsewhere [15].

## METHODS

This review summarizes the available evidence on interventions targeting NCDs in humanitarian emergencies through key examples identified in a larger systematic review [15]. The study is registered at PROSPERO (CRD42018088769).

### Information sources and search strategy

This systematic review follows the reporting guidelines as set out in the Preferred Reporting Items for Systematic Reviews and Meta-Analyses (PRISMA) statement [16]. An experienced medical librarian performed a comprehensive search of multiple databases after consultation with the lead authors and a Medical Subject Heading (MeSH) analysis of key terms provided by the research team. A search of Medline (OvidSP), Medline (PubMed, for in-process and non-indexed citations), Social Science Citation Index, and Global Health (EBSCO) databases was undertaken using relevant controlled vocabulary terms and synonymous free text words and phrases to capture the concepts of non-communicable, chronic and non-infectious diseases, and different types of humanitarian emergencies inclusive of natural disasters, armed conflicts, terrorism, and failed states (see Appendix). The original searches were run August 10, 2015 and were subsequently updated as of December 11, 2017.

### Original study selection

Retrieved references were pooled in EndNote and de-duplicated to 4,430 citations. Two separate screeners independently evaluated the titles, abstracts, and full text of the eligible articles, with vetting by a third reviewer. English, Arabic and French language articles were eligible and no date restrictions were applied. Studies reporting on mental health and associated terms were excluded from this review given the existing evidence for mental health interventions in the available literature [17-19] and our own research question which sought to primarily address the leading four NCDs as outlined by the WHO [2]: cardiovascular disease (CVD), cancer, chronic respiratory disease, and diabetes. Studies conducted in high income countries (HICs), as defined by the 2015 World Bank Country Classification by Income [20], and review articles were excluded. No other restrictions on study type were applied in the original search.

### Intervention study selection

We present here the results of interventions identified as a result of this review, and their effects on populations organized by phase of emergency management as defined above [9]. Publications reporting on interventions targeting NCDs during disasters in LMICs were included if they incorporated core intervention components as defined by the United States (US) Department of Health and Human Services [21]. When evidence-based interventions are scaled-up or reproduced it is critical to identify not only whether an intervention is successful, but also which program elements are essential in making the intervention effective [21]. Presented here are only those interventions identified by our systematic review that featured a description and specification of core intervention components defined by US Department of Health and Human Services as:

> “the context of the program; the core components; the active ingredients to operationally define the core components so they can be taught and learned and can be implemented in typical settings; and a practical strategy for assessing the behaviors and practices that reflect the program’s values and principles, as well as the program’s active ingredients and activities [21].”

### Analysis

Data were extracted into a prespecified data extraction table on the following: (i) study authors and publication date, (ii) study type and design, (iii) geographic location, (iv) NCD addressed by the intervention, (v) target population, (vi) sample size, (vii) crisis type, (viii) phase of emergency management (mitigation, preparedness, response, and recovery), (ix) study duration, (x) key actors and intervention implementers, (xi) description and components of the intervention, (xii) outcomes measured, (xiii) results, and (xiv) author conclusions. A quantitative meta-analysis was not possible due to the limited number of studies and the heterogeneity in study interventions and outcomes, instead a narrative synthesis of results was undertaken.

### Quality Assessment

A quality assessment was conducted using the Newcastle-Ottawa Quality Assessment Scale (NOS) version for cohort studies used for the observational studies [22]. This was selected as it represents a commonly utilized tool for quality assessment of observational studies with demonstrated validity and reliability endorsed by Cochrane reviews [22-24].

## RESULTS

### Search results

In our study we found limited evidence on interventions for NCDs in humanitarian crises (See Table 1). After de-duplication of records, searches identified a total of 4,430 references. 4,342 studies were excluded by title or abstract and 158 articles were read in full. While 85 articles were eligible for the full systematic review, presenting epidemiologic evidence for the burden of NCDs in humanitarian settings, only seven articles described interventions meeting inclusion criteria, as described in the methods section above, and were included in the qualitative synthesis [25-31]. The flowchart per PRISMA is presented in **Figure 1**.

**Figure 1:** PRISMA Diagram

### Characteristics of included studies

Details of each intervention and key outcome measures, study results, and specific study conclusions are presented in **Table 1**. Diseases represented included diabetes mellitus (DM) [25, 26, 28, 29, 31], cancer [27, 30], cardiovascular disorders (CVD) [26, 29, 30], hypertension (HTN) [26], and neuropsychiatric disorders [30]. No studies described interventions addressing chronic pulmonary diseases [i.e., asthma, chronic obstructive pulmonary disease (COPD), etc.] or heart failure. Only two of the seven publications described interventions inclusive of pediatric populations [25, 30].

#### Study design and length

All seven included studies used observational study designs [25-31]. One study was a retrospective review [27]; two were case studies [25, 26], three were cross sectional [28, 30, 31], and one was a community intervention study [29]. Intervention duration ranged from a single health education session [29] to 12 months [30].

#### Quality Assessment

The quality assessment identified several common limitations primarily related to follow-up and comparability (assessed using the Newcastle-Ottawa Quality Assessment Scale). No included study had a defined comparison group or unexposed cohort. Only one observational study reported follow-up procedures for participants [29], whereas the majority of studies did not describe follow-up procedures [25, 27, 30]. The longest follow-up period was six months [26, 29], with the remainder of studies failing to report length of follow-up. Outcome assessment was problematic in several studies which utilized self-reporting of outcomes [25, 26] or a limited scope of intervention assessments measures (number of diabetic kits used without patient outcomes, pre- and post-knowledge tests for community health workers without assessing community impact, etc.) [25, 26, 31]. Assessment of the quality of included studies is provided in tabular format in **Table 2**.

#### Study settings

All seven studies were published between 2007 and 2016; one article was undertaken in the African region [25], two in the Eastern Mediterranean Region (EMRO) [26, 30], three in the Western Pacific region [27, 28, 31], and one in Europe [29]. Two studies reported on interventions taking place in the context of a natural disaster [27, 28]. Four studies reported on interventions taking place in the context of a non-international armed conflict [25, 26, 30, 31] and one study reported on an intervention taking place in the context of an international armed conflict [29]. While five studies reported on the response phase [25-28, 30], and two studies reported on the recovery phase of emergency management [29, 31], no studies reported on interventions conducted in the mitigation or preparedness phases of disaster. Studies evaluating interventions for NCD risk factors were only carried out in the recovery phase of emergency management [29, 31]. Results are presented by phase of emergency management.

### Narrative synthesis by phase of emergency management

#### Response Phase

Multiple articles attempted to shed light on best practices for conducting interventions addressing populations with NCDs during the response phase of emergency management [25-28, 30].

##### Key Interventions conducted during armed conflicts

The study by Besancon et al tested an intervention for patients with DM during an active non-international armed conflict in Mali [25] spearheaded by a local non-governmental organization (NGO) called *Santé Diabète*. The invention’s target population included three subpopulations within Mali: persons residing in active conflict regions, IDPs, and populations housing IDPs, each with distinct barriers to intervention delivery. Both adult and pediatric populations were included.

Significant barriers to chronic diabetes care existed in areas of active conflict including limited DM-related services provided by humanitarian actors, fleeing healthcare workers, a lack of accurate and credible health information among target populations, a focus by traditional humanitarian actors on communicable disease epidemics, interruption of supply delivery, destruction of existing infrastructure, and an absence of local Malian government authority. For IDPs and regions housing IDPs, prominent barriers included lack of supplies at facilities for persons with diabetes already being managed, an additional burden on existing services due to the influx of IDPs, lack of capacity to collect regular data in addition to supplementary data to manage the crisis, limited availability of free health services, and insufficient number of healthcare workers to serve the extensive internally displaced population.

Their intervention included amongst its core components access to medicines, testing equipment, kits for the management of diabetic comas (consisting of insulin, rapid acting insulin, glucose strips, urine test streps, lactated ringer’s solution, normal saline, urinary catheters, etc.), and therapeutics for diabetic foot complications (i.e., antibiotics, Dakin’s solution, compresses, and bandages). These were distributed with data collection sheets to account for displacement of persons and to evaluate utilization of the provided medicines and medical equipment. Telemedical support from specialists at the Hôpital du Mali was available for health professionals practicing in regions most heavily affected by conflict as well as for regions experiencing the largest influxes of displaced persons. Outcomes assessed included the number of emergency kits used for treatment (32 diabetic foot complications and 15 diabetic comas) and number of persons in active conflict regions receiving medications (1,814 persons). For children with Type 1 diabetes, Santé Diabète sponsored immediate evacuation from active conflict areas due to a perceived complexity in managing pediatric diabetes.

The study by Kallab et al [26] assessed an intervention implemented by *Help Age International* conducted in eight outpatient health facilities amongst Syrian refugees and vulnerable Lebanese aged 40 and above with either HTN or type 2 DM during an active non-international armed conflict in Lebanon. Following a baseline needs assessment and pilot phase, a comprehensive portfolio of services covering prevention and management of DM and HTN was provided. Key intervention components included onsite laboratory tests and free of charge medicines distributed on a monthly or quarterly basis to minimize conflict-imposed transportation and financial barriers. Healthcare staff involved in the direct implementation of project services received training on the management of HTN and DM by the Lebanese Cardiology Society and the Lebanese Diabetes Society respectively and patient education was offered in health centers using three modalities; (i) one-on-one during patient enrollment, (ii) informal awareness sessions offered in waiting areas, and (iii) biweekly formal sessions.

The principal barriers to providing diabetic management care in active conflict were: (i) insecurity leading to temporary suspension of humanitarian operations; (ii) the fluid movement of refugees which caused difficulty in accurately assessing and meeting the needs of this population (e.g., following refugee influxes, there were often greater demands for medications than prearranged in a particular region but difficulty in supplying them due to military checkpoints), (iii) limited opening hours of the centers, (iv) transportation costs, (v) medication shortages, (vi) financial barriers to diabetic nutritional goals targets, and (vii) limited knowledge of HTN and DM management amongst healthcare staff [26]. Outcomes assessed included exit interviews (59 beneficiaries), patient follow up, and patient weight reduction. While high patient satisfaction was reported, the study found poor patient follow-up (10% of enrolled participants attended three medical visits) and limited post-intervention weight reduction.

##### Key Interventions conducted during natural disasters

Two articles addressing response to natural disasters identified a need for surgical teams providing humanitarian aid to prepare for comprehensive NCD management in addition to management of soft tissue injuries and orthopedic surgery/ amputations [27, 28]. Read et al [28] described an intervention implemented by a foreign surgical team in the aftermath of Typhoon Haiyan in the Philippines where sepsis from foot injuries in diabetic patients constituted an unexpected majority of the workload (33.3% of all procedures were performed on patients with type 2 DM). While the surgical team stocked metformin, they reported shortages of insulin and sulfonylureas to meet the needs of the high volume of patients requiring these medications. Outcomes were evaluated using a Microsoft Excel database throughout the deployment, recording patient demographics, relationship to the typhoon, and surgical procedure performed.

Marom et al [27] described clinical and ethical dilemmas faced when carrying out surgical interventions for patients with head and neck cancers presenting to a joint Israeli-Filipino field hospital during the subacute period following a 2013 typhoon in the Philippines. Awareness of the specific cancer burden in their country of operation prior to deployment guided the Israeli team’s clinical management decisions, in 1) promoting a lower threshold to perform ultrasound guided fine needle aspiration procedures (FNAs) for diagnosis of thyroid cancer due to the endemic nature of thyroid cancers and goiters in the Philippines and in 2) guiding physical examinations to intentionally examine for cervical nodal metastases even in small tumors due to the over 70% of Filipino head and neck cancers with regional lymph node involvement at presentation [27]. The team found that surgical interventions in patients with advanced head and neck tumors could be performed for therapeutic, palliative, and diagnostic purposes in the setting of a relief mission in cooperation with local health services. However, the team did not perform any reconstructive or debulking procedures in patients who were not likely to benefit from them, often due to advanced disease or lack of long-term follow up. The team noted that their primary goal was to support and maintain regular medical care for the local population in a time of strained resources rather than offer new services which were not present locally at baseline pre-disaster [27]. Outcomes were evaluated using a retrospective review of all charts for patients presenting with head and neck cancers, recording patient demographics, diagnosis, and surgical intervention performed. Ethical dilemmas surrounding the decision of whether to preform diagnostic/therapeutic surgical interventions were highlighted in four key patients, two patients in whom diagnostic/therapeutic surgical interventions were performed and two patients in which the team opted to defer intervention.

#### Recovery Phase

Two articles evaluated interventions for NCD risk factors during the recovery phase of emergency management [29, 31]. Ebling et al [29] found that administration of a single counseling session aimed at lifestyle changes can be effective at decreasing CVD and DM risk factors. Six months after the intervention, the group of 202 returned refugees of war operations in Eastern Slavonia, showed significant weight reduction (28.13% decrease in BMI; p<0.001), increased physical activity (p=0.004), and improved glycemic control (6.09% decrease; p=0.03). However, the intervention had limited success in patients with long standing disease and serum glucose values > 8.5 mmol/L. No control group was evaluated. Similarly, Wagner et al [31] evaluated a diabetes prevention curriculum for community health workers (CHWs) in post-war Cambodia [31]. CHW knowledge of diabetic prevention, assessed by pre- and post-tests, increased significantly after participating in the intervention (p<0.001), however, no assessment of community knowledge retention or patient outcomes post-intervention was provided, significantly limiting the ability to evaluate the intervention’s effectiveness in diabetes prevention.

#### Intervention Financing

Intervention financing in the response phase of emergency management was assessed primarily by McKenzie et al [61] who evaluated cost of care for neuropsychiatric disorders as part of an intervention conducted by the UNHCR. The UNHCR funds tertiary level medical care for refugees based on the cost and acuity of required care by means of application to an Exceptional Care Committee (ECC). Oncologic care exceeded all other disorder category costs (181,815 USD overall; 7,905 USD per applicant). Stroke was costly due to its higher frequency (16%; n=41), while multiple sclerosis was expensive at an individual level (7,502 USD per applicant). Most applications were for emergency care (67%; n□=□176). Of the 20 approved ECC applications for brain tumors, 15% of applications were approved for less than the asked amount —receiving on average only 39% of requested funds. Applications for disc prolapse (50%), cerebral palsy (33%), and tumors (27%) were the most likely to be denied. Six oncology applications were denied due to eligibility, cost, or prognosis. Similarly, Besancon et al [25] reported intervention financing as a major barrier to intervention provision given a lack of response from traditional humanitarian donors during an active non-international armed conflict in Mali and Kallab et al [26] highlighted transportation costs as the primary financial barrier to NCD management amongst Syrian refugees and vulnerable Lebanese during an active non-international armed conflict in Lebanon.

## DISCUSSION

Our review highlights the limited quantity and quality of evidence on the effectiveness of interventions targeting NCDs during humanitarian crises in LMICs. Few articles addressed interventions meeting inclusion criteria [25-31] and no articles addressed interventions conducted in the mitigation or preparedness phases of emergency management, suggestive of a failure to incorporate NCD management into emergency response planning. Commonly cited challenges for providing care in active conflict included: insecurity, the fluid movement of refugees, cost, ethical dilemmas, lack of continuity of care, loss of local healthcare infrastructure and providers, lack of knowledge and experience amongst humanitarian healthcare workers with NCDs, and shortages of medications and medical supplies [25-27, 30].

Successful interventions conducted extensive pre-disaster risk assessments to assess the burden and distinct epidemiology of NCDs amongst affected populations [25, 27, 32-34]; worked in close cooperation with local health services when possible [27]; assessed individual needs of sub-populations in disaster regions (i.e. people still in active conflict regions, IDPs, refugees, and regions housing IDPs); adopted flexibility in the implementation of guidelines and tailoring of activities as per resources and needs of the beneficiaries [25-29]; and promoted task shifting between humanitarian and development actors [25]. Educational background, training and capacity building of staff were found to be essential elements for the success of interventions targeting NCDs due to a perceived lack of experience of healthcare workers in disaster settings with NCDs [26, 31, 35] and successfully allowed for incorporation of community health workers [31]. Strengthening collaboration between humanitarian and development actors to move beyond traditional silos and take on shared roles [36], was also found to promote improved outcomes for NCDs in humanitarian emergencies, building upon development agencies’ experience and long-term funding resources for NCD management in LMICs [25].

Significantly, no studies evaluated interventions targeting NCD risk factors during the mitigation, preparedness, or response phases of emergency management. Accordingly, NCDs were often managed in the these phases of disaster when they presented for emergent management at advanced stages of disease [27, 28, 30] rather than addressed through primary, secondary, or tertiary prevention strategies, as evidenced by McKenzie et al.’s [30] assessment of neuropsychiatric referrals to the UNHCR ECC where 67% of referrals were for emergency care [29, 31]. Delayed presentation of disease is not only more likely to confer worse prognosis for individuals in disaster settings but is also associated with higher cost [30, 37]. This may be addressed by introducing NCD risk reduction interventions in earlier phases of emergency management rather than in the recovery phase of disaster [29, 31]. In highlighting that administration of a single counseling session aimed at lifestyle changes can be effective at decreasing CVD and DM risk factors, the intervention studied in Ebling et al demonstrates that interventions aimed at NCD risk reduction may prove feasible for application in earlier stages of disaster response [29].

Surgical interventions may also provide diagnostic, therapeutic, and palliative options for NCD patients in disaster settings (i.e., diabetic foot amputations, tumor resections, etc.) [27, 28, 38]. Surgical teams deploying to disaster zones should carry NCD diagnostics, equipment, and medications and be appropriately trained in the fundamentals of NCD management [27, 28, 38]. However, teams conducting surgical interventions identified in our review reflected that they deployed to disaster zones unaware and unprepared for a high NCD burden. This was exemplified by Read et al [28] who noted that their surgical team was unaware of such a high proportion of diabetics when planning on delivering a surgical interventions in the disaster setting and recommended as a result of their experience that surgical teams stock a minimum standard of long□ and short□acting insulin, oral hypoglycemics, and glucose testing equipment when deploying to an area where diabetes is prevalent [27, 28, 38].

Finally, oncology patients were found to represent a distinct category in consideration of the ethical, financial, and operational challenges of providing cancer care during an emergency, yet few studies addressed these challenges or proposed evidence-based solutions [27, 30], such as an evaluation of a refugee health financing model for these patients [39]. The oncological interventions identified by our review focused primarily on curative options, rather than addressing palliative care provision, and were often limited to those with a favorable prognosis for treatment [27, 30].

NCDs present a challenge to the traditional humanitarian model based on camp settings of the 20^th^ century where the mainstays of health interventions in humanitarian emergencies were vaccination campaigns, control of infectious communicable disease outbreaks, and nutritional support [12, 13]. Interventions addressing NCDs require humanitarian actors to consider new challenges such as continuity of care, reduction of regular treatment interruptions, cost of care for recurrent or expensive treatments, new diagnostics, specialist physician availability, healthcare provider training, comorbidities, refugee health insurance models, and preventative strategies including regular screening and lifestyle modification [11, 13, 40].

While NCDs have gained increasing attention in the last decade with broader inclusion in the Sphere guidelines [12, 40], the COVID-19 pandemic —as an example of a large scale humanitarian emergency —illuminates the alarming gap in data on NCDs in LMICs [41]. Humanitarian organizations facing the combination of reduced incomes and rising demands due to the pandemic, are reporting plans to scale back critical NCD interventions in 2021, such as hemodialysis for chronic kidney disease patients, in response to substantial funding deficits [42]. Conversely, as NCDs are associated with worse outcomes among those infected with COVID-19, an improved understanding of NCDs has proved critical to a more effective COVID-19 response [41, 43].

### Limitations

There are several limitations to our study. As a result of the limited number of studies evaluating interventions for NCDs during disasters in LMICs, we were unable to draw definitive conclusions from the existing published literature. Only narrative synthesis was used, however alternative methods (meta-analysis, etc.) were not appropriate due to the limited number of studies and the heterogeneity in study interventions and outcomes. Additionally, we may have excluded relevant studies that were not written in English, Arabic, or French due to limited language translation capacity.

## CONCLUSION

We identified only a limited number of interventions designed to address NCDs in humanitarian emergencies, with a particular dearth of studies addressing the mitigation phase of disaster. While several challenges to NCD management such as insecurity and fluid movement of refugees create inherent challenges to NCD management in disasters, the lack of knowledge and training in NCD management amongst healthcare providers as well as the absence of basic medications and supplies for NCD management highlighted in this review are amenable to further intervention. The lack of priority given to patients with NCDs in interventions for humanitarian emergencies requires further study, with significant attention paid to NCD risk factor reduction and the advancement of interventions to address NCDs in the mitigation and preparedness phases of emergency management. Taking immediate action to address these issues is critical given the growing disease burden of NCDs in LMICs experiencing humanitarian crises.

## Data Availability

All data produced in the present work are contained in the manuscript.

## DECLARATIONS

### Availability of data and materials

Data sharing is not applicable to this article as no datasets were generated or analyzed during the current study.

### Authors’ contributions

All authors read and approved the final manuscript.

## Acknowledgements

I thank the anonymous reviewers for their helpful comments.

## LIST OF ABBREVIATIONS

COPD: Chronic Obstructive Pulmonary Disease
CHW: Community Health Worker
CVD: Cardiovascular Disease
DM: Diabetes Mellitus
ECC: Exceptional Care Committee
EMRO: Eastern Mediterranean Region
FNA: Fine Needle Aspiration
HICs: High Income Countries
HTN: Hypertension
IDP: Internally Displaced Person
LMICs: Low and Middle-Income Countries
NCDs: Non-Communicable Diseases
NOS: Newcastle-Ottawa Quality Assessment Scale
MeSH: Medical Subject Heading
NGOs: Non-Governmental Organizations
PRISMA: Preferred Reporting Items for Systematic Reviews and Meta-Analyses
UNHCR: United Nations High Commissioner for Refugees
US: United States
WHO: World Health Organization

## SUPPLEMENTARY FILES

Table 1. Summary of studies examining effectiveness of interventions targeting NCDs during humanitarian crises

Table 2. Quality assessment of observational studies using NOS criteria

Appendix.

