## Supplementary material for "A Review of Interventions for Non-Communicable Diseases in Humanitarian Emergencies in Low-and Middle-Income Countries": Table 1

Table 1. Summary of studies examining effectiveness of interventions targeting NCDs during humanitarian crises

|  | Location | Crisis Type | Phase of emergency management | Intervention duration | Intervention implementers (agency/ies involved) | NCDs addressed | Intervention components | Key actors for intervention implementation (eg CHWs, trained research personnel, established clinic personnel, etc) | Target population | Study type and design (where applicable) | Outcomes Measured (where applicable) | Results (where applicable) | Author/ Study Conclusion |
| --- | --- | --- | --- | --- | --- | --- | --- | --- | --- | --- | --- | --- | --- |
| Besancon (2015)(28) | Region: Africa  Country: Mali | Non-international armed conflict | Response | Unknown Duration (April 2012 start date) | WHO, French Crisis Center, French Development Agency, private donations | Diabetes Mellitus (DM) | 1. Service delivery (support to other NGOs and health facilities; transporting children)  2. Information collection  3. Medical products (development of kits for diabetic coma & skin infections)  4. Financing (covering costs for those in need) | Local HCWs, NGO partners | Malians with diabetes in conflict areas or those displaced. | Case study | Number of people provided with medicines and kits | 1,814 people provided with medicines. 32 kits delivered for diabetic foot complications and 15 for diabetic coma. | People with diabetes should be included as a vulnerable population in the context of emergencies. |
| Kallab (2015)(29) | Region: EMRO  Country of origin: Syria  Host country: Lebanon | Non-international armed conflict | Response | 6 months (November 2014 - May 2015) | HelpAge International (HelpAge), Medecins du Monde (MdM), Amel Association International (AMEL), YMCA Lebanon, American University of Beirut, Centre for Public Health Practice (AUB/CPHP) | Diabetes Mellitus (DM) and Hypertension (HTN) | 1. Provision of services (comprehensive care including promotion, prevention, and management of DM & HTN)  2. Capacity building (training sessions for staff by specialists)  3. Advocacy (for elderly individuals and those with disabilities) | Local HCWs across 8 health facilities (5 health centers & 3 mobile units) | 1,825 Syrian refugees and vulnerable Lebanese aged 40 and above. | Case study | Enrolment | 1,825 patients were enrolled in the program. HTN accounted for 46% of cases, DM 27% of cases, and 27% had both DM & HTN. | NCD management is a fundamental pillar for long-term policy response to the crisis in Syria. |
| Marom (2014)(30) | Region: Western Pacific  Country: Philippines | Natural Disaster | Response | 11 days (November 2013) | Israeli Defense Forces (IDF), Severo Verallo Memorial District Hospital (Philippines) | Cancer: Head and Neck H&N) Tumors | Diagnostic/therapeutic interventions (biopsy, imaging studies, palliative treatment) vs. no intervention | Physicians, Medical Corps professionals, logistic personnel | 1,844 Patients presenting to an integrated Israeli-Filipino medical facility | Retrospective review; case series | Demographics, tumor site, diagnostic studies | Of 1,844 patients, 85 (5%) were found to have H&N tumors. Of those, 68 (80%) were thyroid neoplasms. | In disaster relief, surgical interventions can provide diagnostic, therapeutic, and palliative options for H&N tumors. |
| Ebling (2007)(32) | Region: Europe  Country: Croatia | International armed conflict(35) | Recovery | Single counseling session with 6 months post-intervention follow up (2006) | Dept of Family Medicine, Osijek University School of Medicine; Institute of Public Health for Osijek-Baranya County, Osijek, Croatia | Diabetes Mellitus (DM) & Cardiovascular disorders (CVD) | Single health education session as preventive measure for DM & CVD  -free glucose testing  -healthy lifestyle advice session | Local HCWs, Local Ministry of Health | 202 post-war Returnees to the Osijek Region, Croatia. | Community intervention study | Serum glucose, BMI, eating & lifestyle habits | Significant weight reduction and improvement in glycemic control in 202 respondents six months after intervention | Single counseling session aimed at lifestyle changes can be effective at decreasing CVD risk factors. |
| Read (2015)(31) | Region: Western Pacific  Country: Philippines | Natural Disaster | Response | 21 days (November 2013) | Australian Medical Assistance Team (AUSMAT), Save the Children UK | Diabetes Mellitus (DM) | Surgical procedures (i.e. soft tissue, amputation, orthopedic, abdominal | 37 personnel (30 medical and nursing, 7 logistics); including 2 surgeons, 2 anesthesiologists, 1 emergency physician, nurses | 131 Filipinos with surgical needs after Typhoon Haiyan | Cross Sectional | Injuries, Surgery type, Diabetes status | 222 surgical procedures performed in 131 patients; 30 with DM (22.9%) requiring 74 procedures (33.3%) on foot wounds. | Recommend ability to measure glucose and deliver insulin when deploying to disaster areas where DM prevalent. |
| McKenzie (2015)(33) | Region: EMRO  Country of Asylum:  Jordan  Country of Origin:  Iraq, Syria | Non-international armed conflict | Response | 12 months (2012-2013) | United Nations High Commissioner of Refugees (UNHCR | Neuropsychiatric disorders including Multiple Sclerosis, Stroke, and Central Nervous System(CNS) Tumors | Exceptional Care Committee determination for eligibility to receive UNHCR funded tertiary care | UNHCR funded tertiary care for refugees based on the cost and immediacy of care required via application to an Exceptional Care Committee (ECC) | 223 Syrian and Iraqi refugees residing in Jordan | Cross sectional | Demographics, cost of intervention, diagnoses | Neuropsychiatric applications accounted for 11% (264/2526) of all ECC applications. The most expensive care per person was for brain tumor (7,905 USD), multiple sclerosis (7,502 USD), and nervous system trauma (6,466 USD), although stroke was the most frequent diagnosis. | Investment in primary and public health initiatives for neuropsychiatric disorders may reduce the demand for costly, exceptional, and emergency care amongst refugees. |
| Wagner (2016)(34) | Region: Western Pacific  Country: Cambodia | Non-international armed conflict | Recovery | 6 months (unknown intervention dates) | Khmer Health Advocates | Diabetes Mellitus (DM) | Diabetes training program for Cambodian community health workers (CHWs)  to deliver diabetes  prevention in their villages. | Cambodian Diabetes Association and Trained Health Center chiefs | 185 local CHWs. Study did  not address the efficacy of the intervention in modifying villagers’  risk for diabetes | Cross sectional | Pre-test and post-test evaluation of CHW knowledge. Study did  not address the efficacy of the intervention in modifying villagers’  risk for diabetes. | After an initial evaluation with a pre-test and post-test, community health workers received continuing education and were instructed to teach the content they learned to the people in their villages. Knowledge scores increased significantly after teaching the classes (p<0.001). | Community health workers were able to effectively learn a diabetes prevention curriculum suggesting that they would be effective at disseminating the information |
