## Supplementary material for "A Review of Interventions for Non-Communicable Diseases in Humanitarian Emergencies in Low-and Middle-Income Countries": Table 2

**Table 2. Quality assessment of observational studies using NOS criteria**

| **Author, Date [Ref)** | **Selection** | | | | **Comparability** | **Outcome** | | |
| --- | --- | --- | --- | --- | --- | --- | --- | --- |
|  | **Representativeness of the exposed cohort** | **Selection of the non-exposed cohort** | **Ascertainment of exposure** | **Demonstration that outcome of interest was not present at start of study** | **Based on design and analysis** | **Assessment of outcome** | **Was follow-up long enough for outcomes to occur** | **Adequacy of follow up of cohorts** |
| Besancon (2015)(28) | **D**  No description given | **n/a**  No comparison group | **B***  Interviews and clinical assessmen**t** | **B**  Not stated | **n/a**  No comparison group | **C**  Self-report | **B**  Length undefined | **D**  No statement |
| Ebling (2007)(32) | **A***  Randomized sample selected from returned refugees | **n/a**  No comparison group | **B***  Interviews and clinical assessmen**t** | **A***  Outcomes focussed on changes from baseline | **n/a**  No comparison group | **B***  Record linkage to EMR database | **A***  6 months | **A***  Complete follow up - all subjects accounted for |
| Kallab (2015)(29) | **A***  Patients with DM or HTN who seek care | **n/a**  No comparison group | **A***  EMR record and clinical assessment | **B**  Not stated | **n/a**  No comparison group | **C**  Self-report | **A***  6 months | **D**  Statement inadequate |
| Marom (2014)(30) | **A***  Patients with head and neck tumours who seek care | **n/a**  No comparison group | **A***  Prior clinical records and clinical screening | **A***  Outcomes focussed on changes from baseline | **n/a**  No comparison group | **B***  Record linkage to EMR database | **B**  Length undefined. Follow-up with local staff members | **D**  No statement |
| Read (2015)(31) | **A***  Patients with surgical disease management who seek care | **n/a**  No comparison group | **A***  EMR record | **B**  Not stated | **n/a**  No comparison group | **B***  Record linkage to EMR database | **B**  Length undefined | **D**  No statement |
| McKenzie (2015)(33) | **A***  Patients with neuropsychiatric disease who seek care | **n/a**  No comparison group | **A***  EMR record | **B**  Not stated | **n/a**  No comparison group | **B***  Record linkage to EMR database | **B**  Length undefined | **D**  No statement |
| Wagner (2016)(34) | **C**  Selected group of community health workers | **n/a**  No comparison group | **A***  Secure record -post-test | **A***  Outcomes focussed on changes from baseline | **n/a**  No comparison group | **B***  Record linkage | **B**  Length undefined | **D**  Statement inadequate |
| Quality assessment per NOS criteria for cohort studies. Stars (*) awarded if study reached threshold of high quality for that category. Letters and descriptions given to cross-reference with NOS coding manual. For further information on NOS see <http://www.ohri.ca/programs/clinical_epidemiology/oxford.asp>. For the NOS manual see <http://www.ohri.ca/programs/clinical_epidemiology/nos_manual.pdf>, and for the scoring scale see <http://www.ohri.ca/programs/clinical_epidemiology/nosgen.pdf> | | | | | | | | |
