## Supplementary material for "A Review of Interventions for Non-Communicable Diseases in Humanitarian Emergencies in Low-and Middle-Income Countries": Figure 1

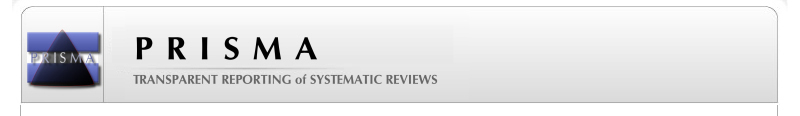
**PRISMA 2009 Flow Diagram**

**Screening**

**Included**

**Eligibility**

**Identification**

Records identified through database searching after duplicates removed
(n =4430)

SSCI=1379

Medline/Ovid=1690

PubMed=673

Global Health=88

Records screened
(n = 4430)

Records excluded
(n =4342)

Full-text articles assessed for eligibility
(n = 158)

Full-text articles excluded, with reasons
(n = 73)

Not in a disaster setting

Study did not take place in	an LMIC

Study did not include information pertaining to NCDs

Study methodology was a review article

Studies included in qualitative synthesis as published in the full systematic review
(n = 85)

Studies included in the qualitative synthesis meeting the criteria described for an analysis of an intervention as published in this review

(n=7)
