## appendix for "A Review of Interventions for Non-Communicable Diseases in Humanitarian Emergencies in Low-and Middle-Income Countries"

**Pubmed Search Strategy**

((Noncommunicable OR non-communicable OR "chronic disease*" OR non-infectious OR hypertension OR "high blood pressure" OR diabetes OR "chronic renal insufficiency" OR "myocardial ischemia" OR "coronary artery disease" OR cva OR cancer OR neoplasms OR "chronic obstructive pulmonary disease" OR copd OR "lung disease*" OR "chronic illness" OR myopia* OR astigmatism OR anisometropia OR aniseikonia OR asthma OR "reactive airway disease") AND ("natural disaster*"[tiab] OR famine[tiab] OR flood*[tiab] OR earthquake*[tiab] OR cyclone*[tiab] OR drought*[tiab] OR war[tiab] OR wars[tiab] OR tornado*[tiab] OR hurricane*[tiab] OR tsunami* OR torture[tiab] OR terrorism OR "conflict zone*" OR genocide OR "ethnic cleansing" OR "economic collapse" OR "economic disaster" OR "tropical storm*" OR "tidal wave*" OR typhoon* OR landslide* OR volcano*[tiab]) AND ((English[lang] OR Arabic[lang] OR French[lang]))) OR ((Noncommunicable OR non-communicable OR "chronic disease*" OR non-infectious OR hypertension OR "high blood pressure" OR diabetes OR "chronic renal insufficiency" OR "myocardial ischemia" OR "coronary artery disease" OR cva OR cancer OR neoplasms OR "chronic obstructive pulmonary disease" OR copd OR "lung disease*" OR "chronic illness" OR myopia* OR astigmatism OR anisometropia OR aniseikonia OR asthma OR "reactive airway disease") AND (Refugee* OR "asylum seeker*" OR transients OR "displaced person*" OR "displaced people*")) Filters: English; Arabic; French
